# KNOWLEDGE, ATTITUDE AND PRACTICE OF YOUNG WOMEN (AGE 18-25) IN BANGALORE CITY WITH REGARDS TO SELF-BREAST EXAMINATION

**DOI:** 10.1101/2024.06.26.24309399

**Authors:** Swastik Pandita, Andrea Dias, Shireen Laturkar, K A Shashank

## Abstract

**Introduction:** Breast cancer is a global health concern and a leading cause of morbidity and mortality. Breast self examination (BSE) is a commonly recommended screening method for breast cancer. This study intends to assess the knowledge of breast cancer and its screening in women aged 18-25 in Bangalore city and to assess the knowledge, attitude and practice of women aged 18-25 in Bangalore city with regards to breast self examination among the female college students studying non medical courses in Bangalore City, Karnataka, India.

**Materials and methods:** A questionnaire based study was conducted among women aged 18-25 in Bangalore city and their scores in the fields of knowledge, attitude, and practice were calculated.Independent sample t-test and one-way analysis of variance (ANOVA) test of significance were used to examine the relation between the demographic characteristics and scores.

**Results:** The mean knowledge score was 3.05 (SD:1.37, range:0-6). Knowledge scores significantly differed across family income per month (P<0.05), age groups (P<0.001), and level of education of father (P<0.05). The mean attitude score was 3.95 (SD:1.78, range:1-8). The mean practice score was 0.83 (SD: 0.954, range:0-3).Practice scores varied significantly across age groups (P<0.05), level of education of mother (P<0.05) and occupation of father (P<0.001).The correlation coefficients are 0.484 and 0.257 respectively (p<0.05).

**Conclusion:** Breast Self Exam (BSE) is an easy method of early detection of breast cancer. There is a need to develop and adopt culturally appropriate and proven interventions to inform and train women about BSE in a diverse country like India. The study can be used as a basis to help raise awareness about BSE and its benefits among the same.

## INTRODUCTION

Breast cancer is a global health concern and a leading cause of morbidity and mortality among women.^1^ It is the second most common cancer in the world and the most frequent cancer among women.^2^ In 2018, there were 2.1 million newly diagnosed breast cancer cases among women, accounting for almost one in four cancer cases among women.^3^ Breast self examination (BSE) is a commonly recommended screening method.^4^ During BSE, a woman can simply look and feel each of her breasts for the presence of lump, swelling, distortion or any other significant change in the area of her breast. It is a simple and effective practice which does not require clinical expertise or any particular equipment. It increases the chances for early treatment and as a consequence, the survival rate in women.^5^ The American Cancer Society recommends BSE for early detection of breast cancer as it assists women to become familiar with the appearance and sense of their breasts, and helps them to detect changes in their breasts as soon as possible. In resource constrained settings BSE has been reported to be culturally and religiously acceptable, friendly and incurring no cost.^6^ An effective way of disseminating knowledge about breast cancer is by educating adolescent women regarding the same. In developing countries like India, Simply trumpeting screening as a solution without an adequate diagnostic and therapeutic infrastructure to support it is bound to fail. It requires a large co-ordinated information campaign to reach the target population, infrastructure development of suitable facilities and, most importantly, a large cohort of adequately trained health practitioners to deal competently with the resulting workload. If combined with accessible and effective treatment facilities, it is surely helpful in early detection and better course of action.Therefore, this study intends to assess the knowledge of breast cancer and its screening in women aged 18-25 in Bangalore city and to assess the knowledge, attitude and practice of women aged 18-25 in Bangalore city with regards to breast self examination among the female college students studying non medical courses in Bangalore City, Karnataka, India.

## Materials and methods

### Design and settings

To measure the KAP towards breast self-examination of the women aged 18-25 in Bangalore city, a web-based questionnaire was developed for this study and used for data. The questionnaire was posted on social media platforms (Facebook and WhatsApp). Privacy of the original post was set as “public” to enable more people to participate in the questionnaire.

### Participants Inclusion Criteria

Women studying or working in non-medicine or non-healthcare fields who were aged 18-25 years and resided in the city of Bangalore, India were eligible for the study.

### Exclusion Criteria

Women who were studying or working in medicine and allied fields and eligible women who did not give prior consent were excluded from the study.

### Informed written consent

Will be obtained in writing prior to administering the questionnaire.

### Sample Size

#### Sample Size calculation

1. Estimate of the expected proportion (p) of knowledge of breast self-examination among market women =0.8
2. Desired level of absolute precision (d) = 0.08

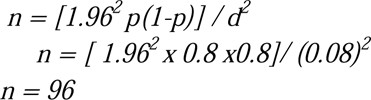

3 Assuming 4% will declining to participate in the study Minimum Sample size = 96 + 4 = 100 participants

### Study tools

After reviewing the literature, a self-administered questionnaire was designed by the researchers to collect the necessary data. The questionnaire was divided into four parts; the first part consists of 10 questions and contains sociodemographic characteristics, age, address, economic status,occupation of father,occupation of mother, level of education of participant, father and family history of breast cancer or breast-related diseases.

The second part contains 11 questions which aim at assessing the knowledge of the participant regarding BSE, breast cancer and mammography. The questions were based on risk factors, signs and symptoms, methods of screening, methods of early detection and diagnosis of Breast cancer and knowledge of performing BSE.

The third part contains 8 questions regarding the attitude of the participant towards BSE, willingness to perform BSE regularly,the factors which may hinder them from doing so and whether they would consult a doctor in case of any breast abnormalities or not.

The fourth part contains 4 questions regarding the practices of the participant with regards to self breast examination, on whether or not they have self examined before, frequency of examination, reasons for not examining and method used for examining the breast.

### Statistical Analysis

Independent sample t-test and one-way analysis of variance (ANOVA) test of significance were used to examine the relation between the demographic characteristics and scores. Correlation between knowledge, attitude, and practice was assessed through Pearson’s correlation coefficient. Jamovi^8^ (Version 2.5) based on R: the R project for statistical computing, was used to analyze the data.

The knowledge,attitude and practice score was calculated by recording the questions by giving one point to the correct answer zero to the incorrect answer. Attitude about hindering factors was calculated by recording of the Likert scale, wherein 1 point was allotted was for either “disagree” or “strongly disagree” as all the hindering factors displayed a negative attitude towards BSE.

### Ethical consideration

The present study was approved by the ethics committee at Shri Atal Bihari Vajpayee medical college and research institute, Bangalore, Karnataka, India. The purpose of this research was explained to all participants and were assured of confidentiality by the researcher. The survey also included an informed consent form, which allowed participants to tick to confirm their consent. Completion of this form followed by completion of the survey showed that participants consented to the study. Participation in the study questionnaire was optional. The anonymity, confidentiality and the fact that the study results would be used for research purposes only were explained in the consent form along with the questionnaire.

## RESULTS

TABLE 1: Demographic characteristics of participants, score of BSE knowledge by demographic variables and relation between the demographic characteristics and scores. A Total Number Of 100 Participants Completed The Survey questionnaire. The mean knowledge score was 3.05 (SD:1.37, range:0-6). This corresponds to a 50.83% correct rate on the knowledge test. Knowledge scores significantly differed across family income per month (*P 0.007*), age groups (*P* 0.001), and level of education of father (*P 0.037*) (Table 1). Occupation of mother did not make a significant difference in BSE knowledge. Knowledge regarding breast cancer was the highest (95%) whereas knowledge regarding the ideal time period to perform BSE was the lowest (5%). Furthermore, 66% of participants had heard about breast self examination before, with social media (34%) being their primary source of information regarding BSE. 29% participants knew the correct frequency of performing BSE. only 16% of the participants have ever been taught to perform BSE.

**TABLE 1:**
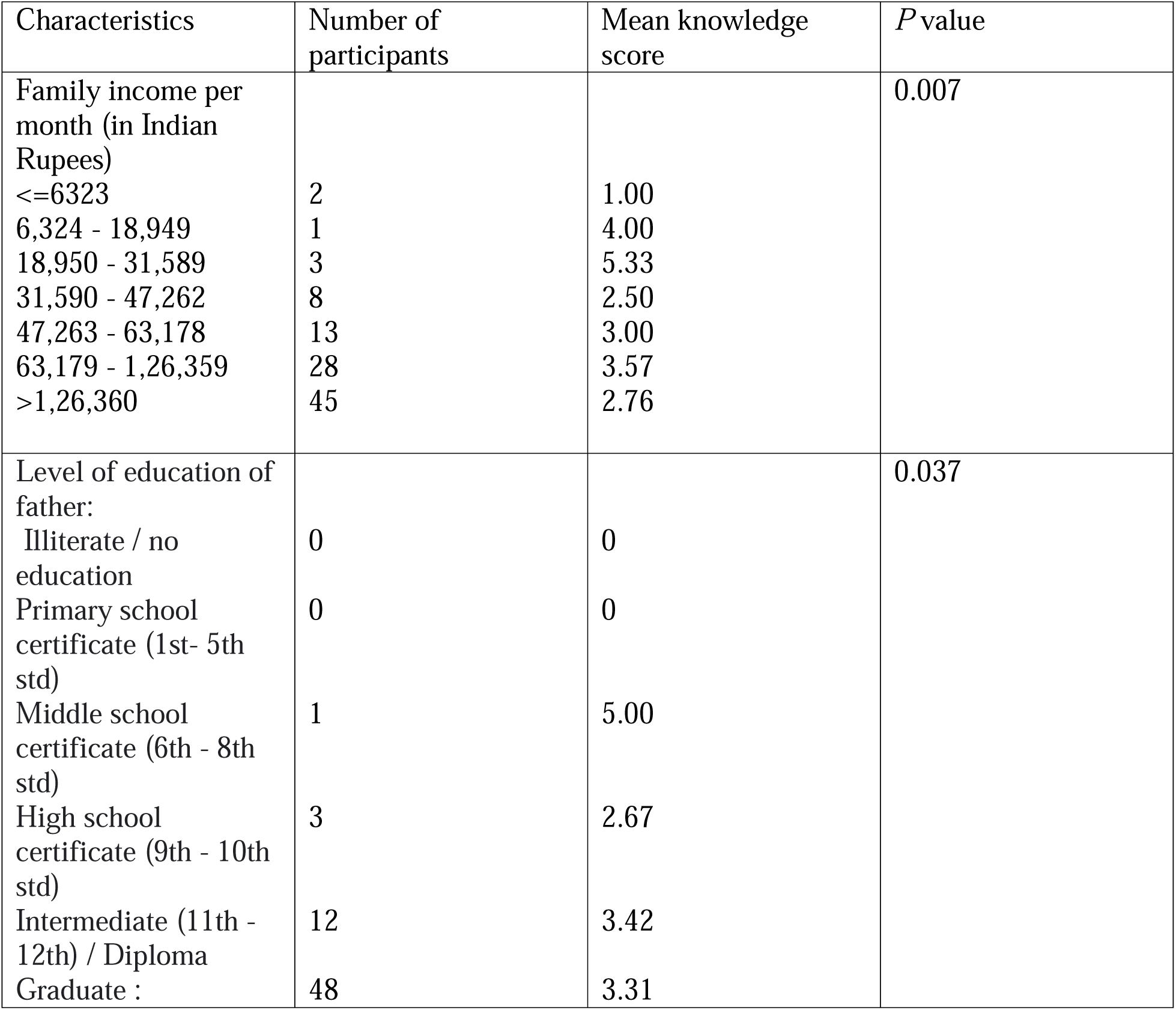
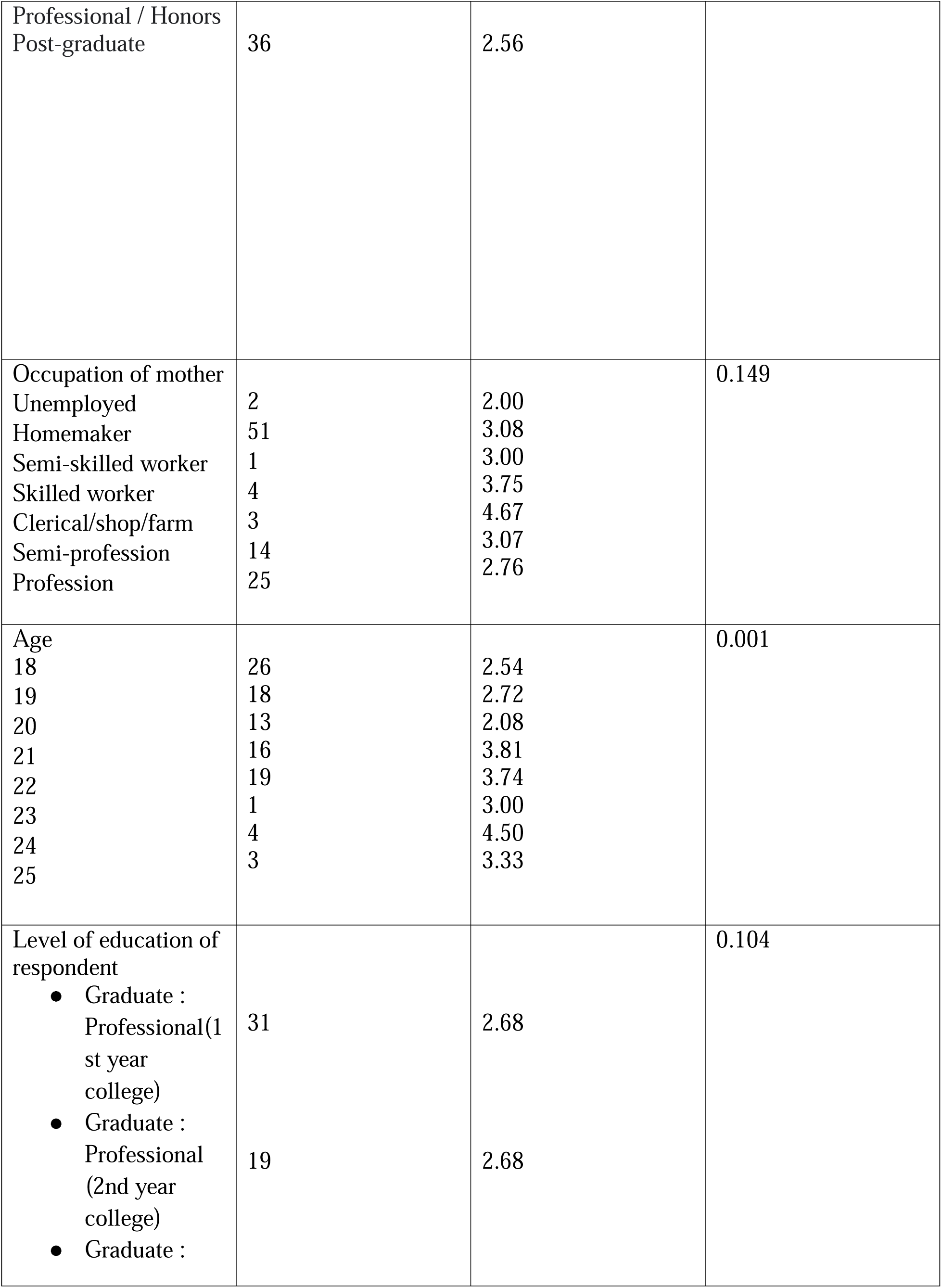
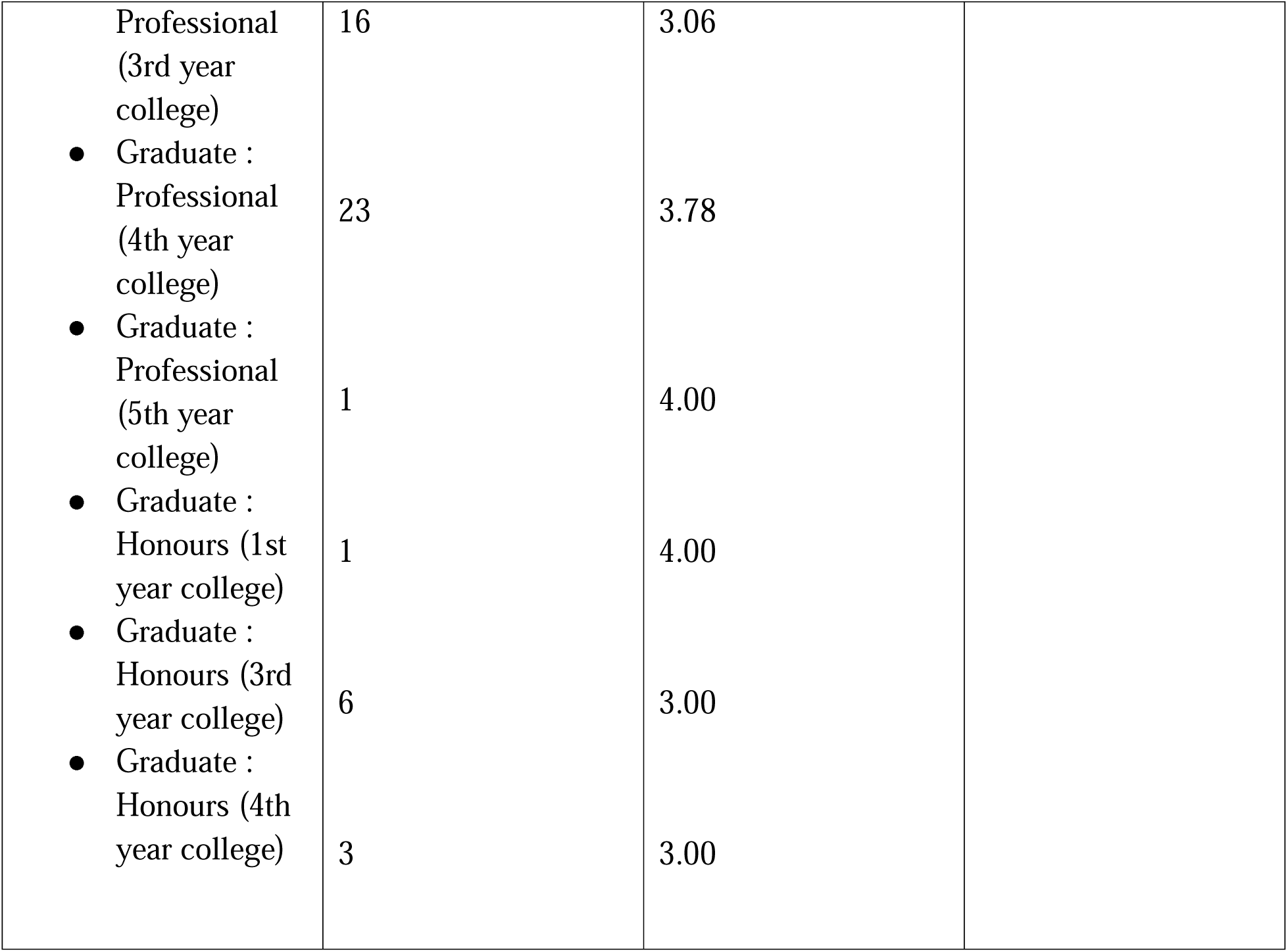
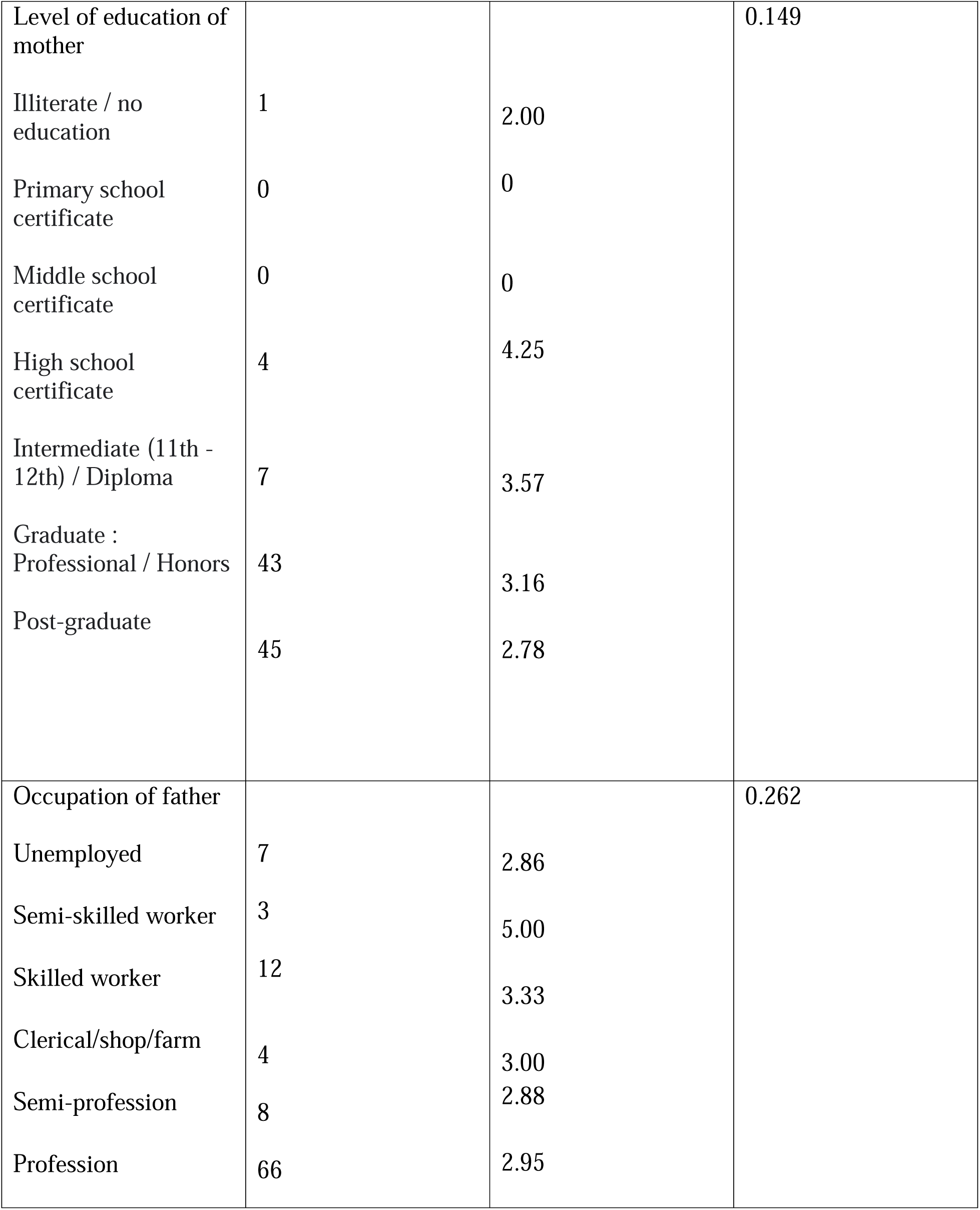
Demographic characteristics of participants, score of BSE knowledge by demographic variables and relation between the demographic characteristics and scores:

Regarding the attitudes, willingness to perform BSE regularly was very high (88%) along with the willingness to consult a doctor (96%). Attitude scores were not significantly different for any demographic variable (table 2). The mean attitude score was 3.95 (SD:1.78, range:1-8). Absence of symptoms was the primary factor hindering the participants regarding performing BSE (73%) along with lack of knowledge regarding BSE (61%).

**Table 2:** Attitudes towards Breast Self-examination by demographic variables.

| Characteristics | Number of participants (%) | Attitude score | *P* value |
| --- | --- | --- | --- |
| Family income per month (in Indian Rupees) |  |  | 0.105 |
| ≤6323 | 2 | 2.00 |  |
| 6,324 - 18,949 | 1 | 4.00 |  |
| 18,950 - 31,589 | 3 | 2.67 |  |
| 31,590 - 47,262 | 8 | 2.88 |  |
| 47,263 - 63,178 | 13 | 4.32 |  |
| 63,179 - 1,26,359 | 28 | 3.91 |  |
| >1,26,360 | 45 |  |  |
| Level of education of father: |  |  | 0.140 |
| Illiterate / no education | 0 | 0 |  |
| Primary school certificate | 0 | 0 |  |
| Middle school certificate | 1 | 4.42 |  |
| High school certificate | 3 | 2.00 |  |
| Intermediate (11th - 12th) / Diploma | 12 | 2.23 |  |
| Graduate : | 48 | 4.21 |  |
| Professional / Honors |  |  |  |
| Post-graduate | 36 | 3.64 |  |
| Occupation of mother |  |  | 0.333 |
| Unemployed | 2 | 1.5 |  |
| Homemaker | 51 | 4.04 |  |
| Semi-skilled worker | 1 | 7.00 |  |
| Skilled worker | 4 | 4.25 |  |
| Clerical/shop/farm | 3 | 3.67 |  |
| Semi-profession | 14 | 3.71 |  |
| Profession | 25 | 3.96 |  |
| Level of education of respondent |  |  | 0.950 |
| ● Graduate : |  |  |  |
| Professional<br>(1st year college) | 31 | 3.94 |  |
| • Graduate :<br>Professional<br>(2nd year college) | 19 | 3.89 |  |
| • Graduate :<br>Professional<br>(3rd year college) | 16 | 4.13 |  |
| • Graduate :<br>Professional<br>(4th year college) | 23 | 4.09 |  |
| • Graduate :<br>Professional<br>(5th year college) | 1 | 5.00 |  |
| • Graduate :<br>Honours (1st year college) | 1 | 4.00 |  |
| • Graduate :<br>Honours (3rd year college) | 6 | 3.50 |  |
| • Graduate :<br>Honours (4th year college) | 3 | 3.00 |  |
| Level of education<br>of mother |  |  | 0.237 |
| Illiterate / no<br>education | 1 | 2.00 |  |
| Primary school<br>certificate | 0 | 0 |  |
| Middle school<br>certificate | 0 | 0 |  |
| High school certificate | 4 | 3.00 |  |
| Intermediate (11th - 12th) / Diploma | 7 | 5.29 |  |
| Graduate : Professional / Honors | 43 | 3.81 |  |
| Post-graduate | 45 | 4.00 |  |
| Occupation of father |  |  | 0.703 |
| Unemployed | 7 | 3.43 |  |
| Semi-skilled worker | 3 | 5.00 |  |
| Skilled worker | 12 | 4.00 |  |
| Clerical/shop/farm | 4 | 3.00 |  |
| Semi-profession | 8 | 3.88 |  |
| Profession | 66 | 4.02 |  |

In addition, the mean practice score was 0.83 (SD: 0.954, range:0-3). Practice scores varied significantly across age groups (*P* 0.039), level of education of mother (*P 0.031*) and occupation of father (*P* 0.001) (table 3). However, the practice was poor as only 35% participants had performed BSE prior, with 59.4% participants citing lack of prior knowledge about the steps and methods of performing BSE. Furthermore, only 45% participants knew the correct methods of performing BSE.

**Table 3:**
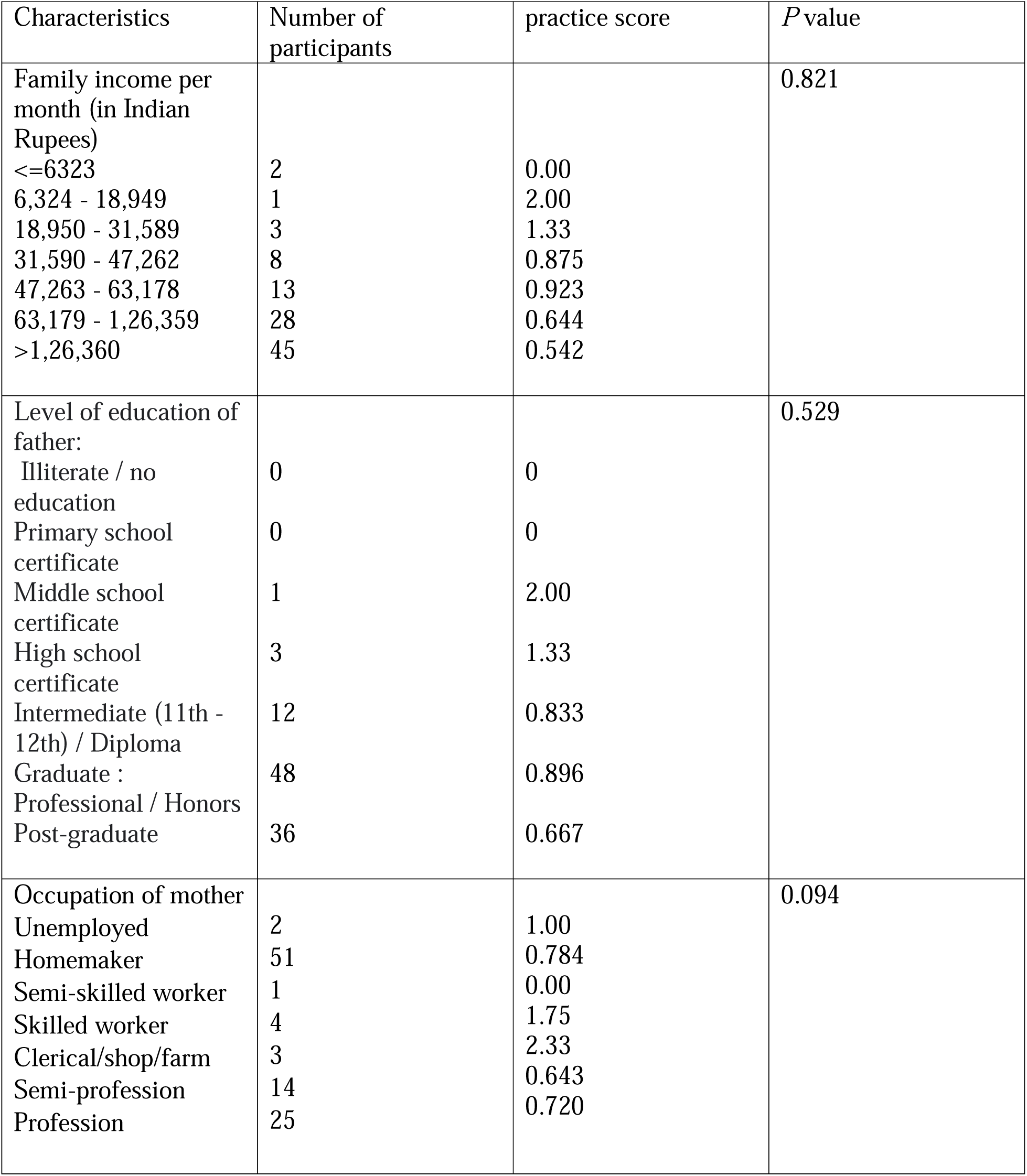
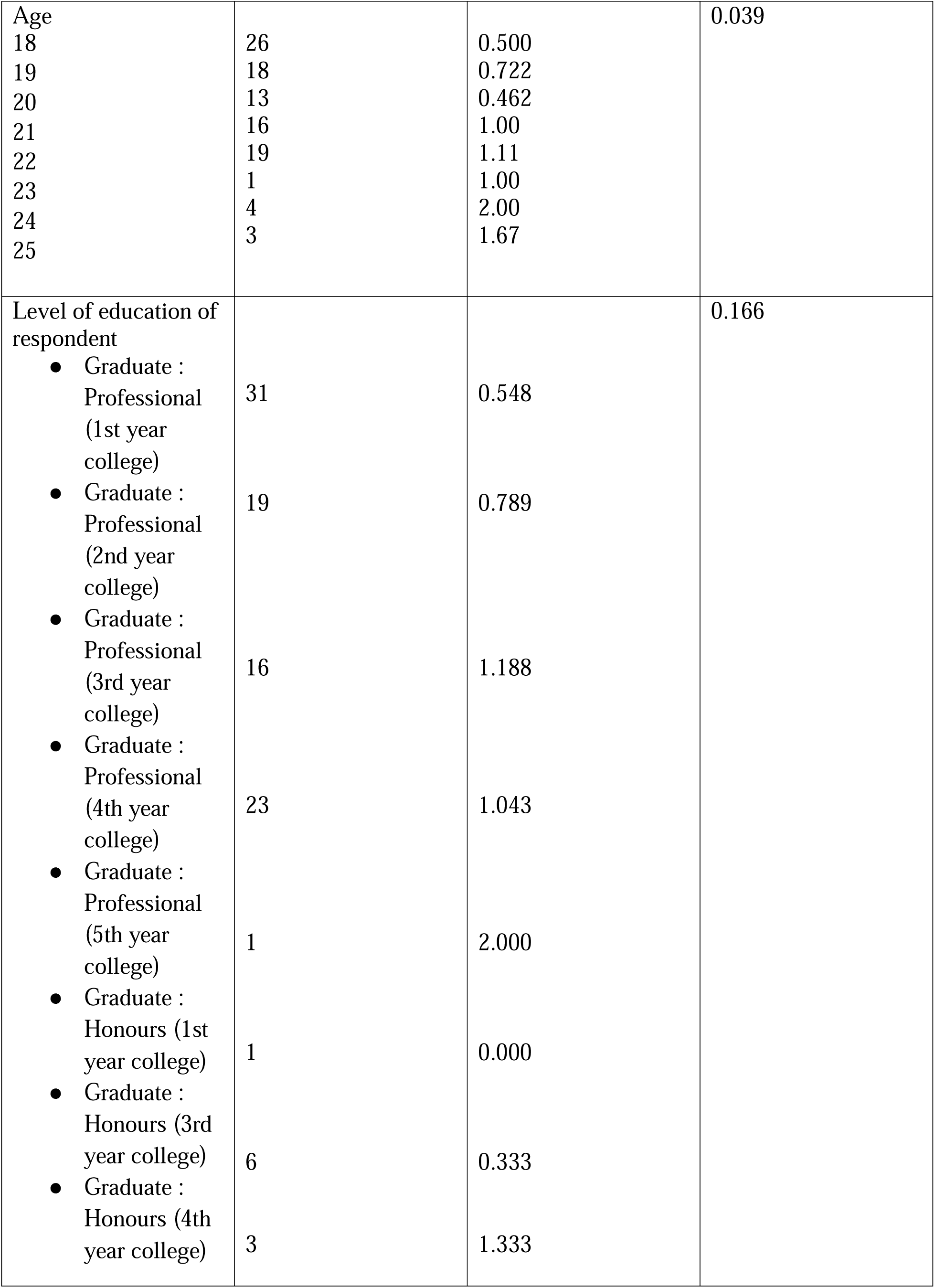
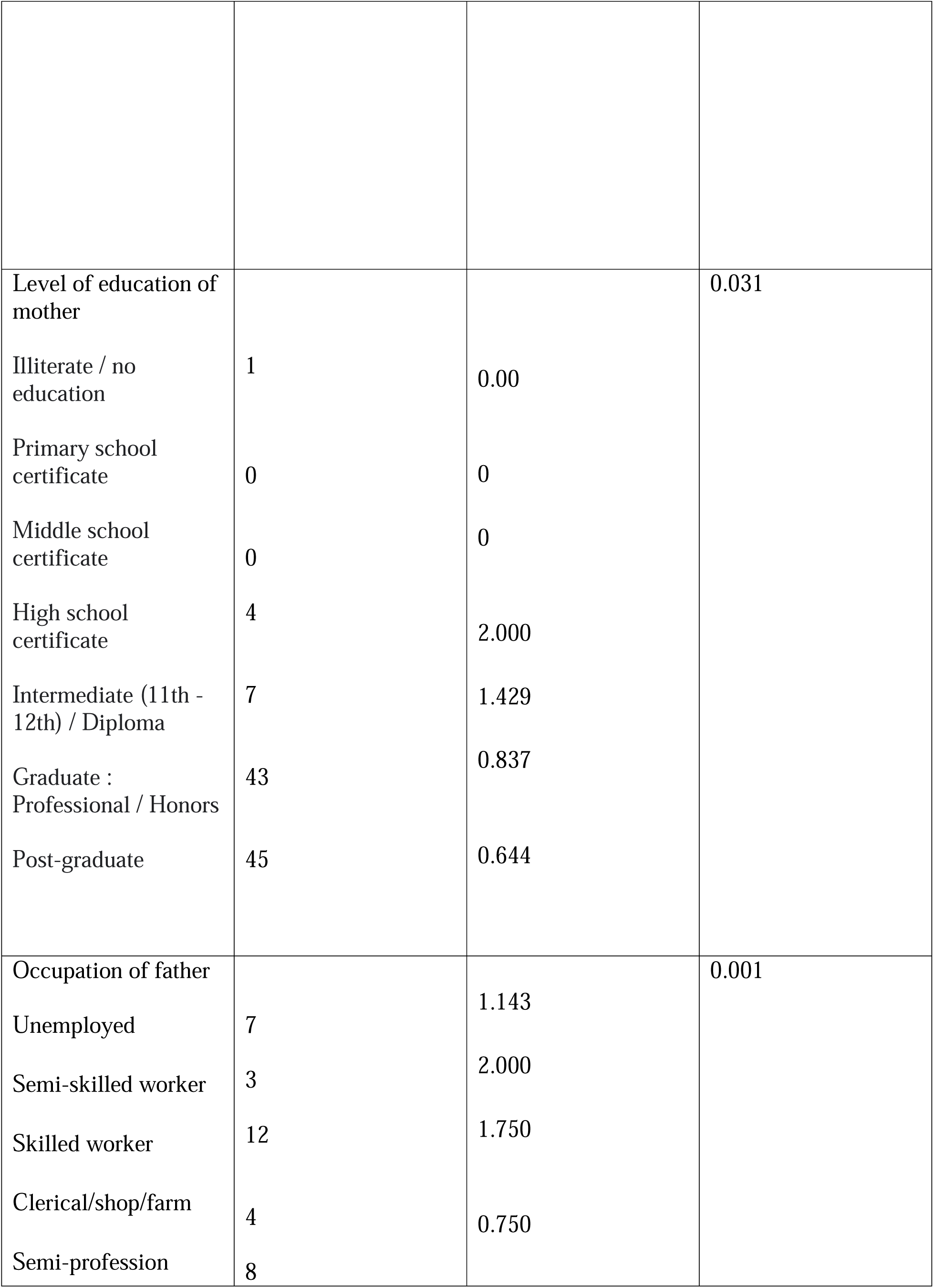
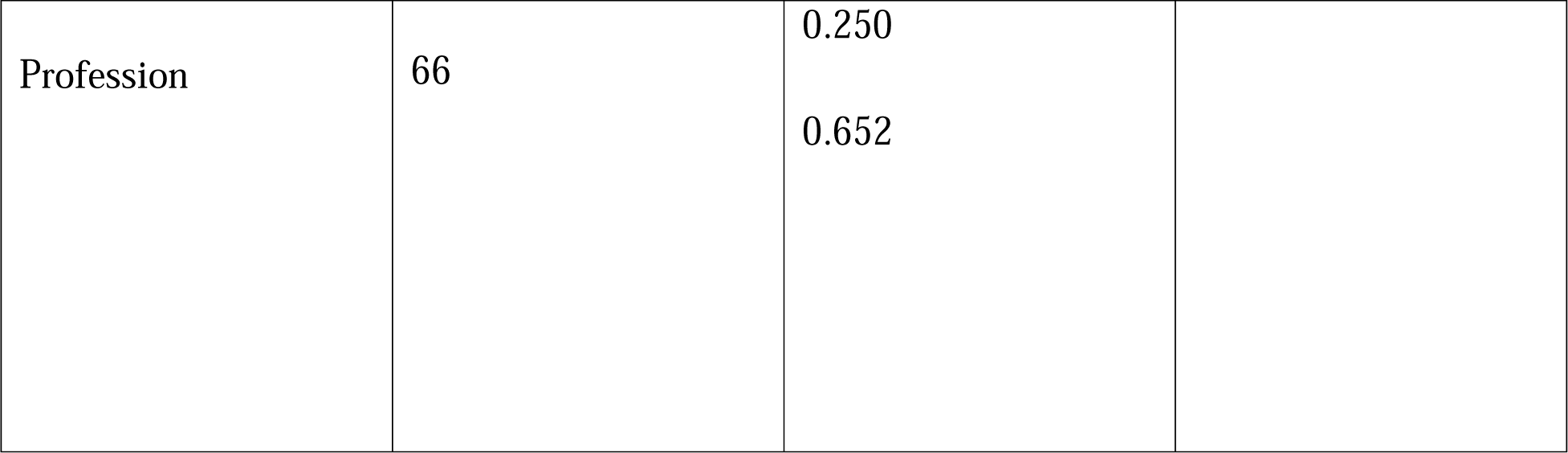
Practice towards Breast Self-examination by demographic variables.

According to the correlation matrix, there was a positive and significant association between knowledge-practice and knowledge-attitudes (table 4). The correlation coefficients are 0.484 and 0.257 respectively.

**Table 4:** Correlation Matrix among knowledge, attitude and practice.

|  |  | **knowledge**<br>**score** | **practice**<br>**score** | **attitude**<br>**score** |
| --- | --- | --- | --- | --- |
| knowledge<br>score | Pearson'<br>s r | — |  |  |
|  | df | — |  |  |
|  | p-value | — |  |  |
| practice<br>score | Pearson'<br>s r | 0.484 | — |  |
|  | df | 98 | — |  |
|  | p-value | <□.001 | — |  |
| attitude score | Pearson'<br>s r | 0.257 | 0.078 | — |
|  | df | 98 | 98 | — |
|  | p-value | 0.010 | 0.440 | — |

## Discussion

Studies conducted in other countries indicate that the need to raise awareness about breast cancer among female university students is likely relevant globally. The survival rate of patients with breast cancer is poor in India as compared to Western countries due to earlier age at onset^9^, late stage of disease at presentation, delayed initiation of definitive management and inadequate/fragmented treatment.

This study is expected to be among the few studies done to examine the KAP towards BSE among the south Indian population. In this population of respondents, the overall correct rate of around 51% on the knowledge questions indicated that most respondents have modest knowledge about BSE. Screening methods are essential in the early detection of BC and lead to decreased morbidity and mortality from cancer.^10^ Starting from the age of 20, every woman should perform BSE regularly.^11^ Similar studies conducted in Nepal^12^ showed about the same proportion of women had modest knowledge (44.3%). Similarly, other studies conducted in Ghana^13^ (43.3%) showed that a similar proportion of respondents had modest knowledge. As far as Indian studies are concerned, a study among female IT professionals in Bangalore,India showed 92% of the respondents having modest knowledge.^14^ This extremely high number might be due to IT professionals having better awareness as they usually fare better on factors like education and incomes. In contrast, a similar study in a rural area of Trichy, India showed appallingly low, only 26% of the respondents were aware of BSE, of which 18% had ever examined their breast, and 5% practiced it regularly.^15^ The current study showed that 35% of women have performed BSE prior. Among the various sources of information regarding BSE, social media (34%) was the most common source of information. The results entice the need for mass media to focus more about breast cancer and BSE to reach all women. A good proportion of participants heard about BSE and knew that it is useful in early detection of breast cancer.This finding is consistent with similar studies performed.^16,17,18^ Only a small percentage (20%) of respondents knew the correct frequency of performing BSE and only 16% of the respondents had ever been taught BSE. This data suggests a knowledge deficit regarding the screening practices and can be a hindrance factor and detrimental in the long run as far as adoption of such practices is concerned.^19^ The unwavering levels of BSE knowledge and awareness could be due to India’s poor public institutional cancer care which, suffers from issues like inefficient administration, poor governance, corruption, lack of resources, etc., has largely lacked organized BSE and breast cancer awareness campaigns. Effective cancer prevention can be achieved through educating the masses about the disease and what they can do to prevent it through effective screening.^20^ Accounts of properly planned awareness campaigns establish their effectiveness.^21^ BSE and other interventions reduce late diagnoses and thus reduce mortality.^22^ Our results further demonstrated that knowledge scores significantly differed across family income per month, age groups, and level of education of the father. Attitude scores were not significantly different for any demographic variable. As per the received responses, the primary factors behind a poor attitude score were lack of knowledge regarding the steps of BSE and absence of any related symptoms. Our results suggest that the practice of the respondents with regards to BSE was poor with only 35% of the respondents having ever performed BSE with the lack of knowledge regarding how to perform BSE being the primary reason (59.4%) and only 45% having the knowledge of various methods used to perform BSE. Our study has few limitations. The use of self-developed questionnaires also introduced bias in the data which is attributable to the phrasing of questions and the relative interpretation of responses. Our data is limited to one urban city of India which largely excludes data from rural villages. In addition,our study was confined to women in non-medical fields. Furthermore, our study is quantitative in nature which inhibits a deeper dive into the responder’s perspective.

## Conclusion

Breast Self Exam (BSE) is an easy method of early detection of breast cancer. There is also the need to develop and adopt culturally appropriate and proven interventions to inform and train women about BSE in a diverse country like India. Taught early, BSE could prove to be a powerful tool in aiding the prevention and control of breast cancer among Indian Women where prevalence and mortality are higher. Understanding and assessing the existing knowledge and current attitude and practice of young women in India is the first step in working towards this goal. The above study only aims to find the knowledge and attitude of young women aged 18-25 regarding breast cancer and self breast examination. The information generated from the study can be used for further research on a more diverse study population which includes and is not restricted to women of various age groups. The study can also be used as a basis to help raise awareness about BSE and its benefits among the same. Further, practical studies involving demonstration of BSE followed by assessment could be conducted.

## Abbreviations

BSE: Breast self-examination
CA: Carcinoma
SD: Standard deviation
KAP: Knowledge, attitude, practice

## Glossary

breast self-examination: A breast self-exam is an early detection tool that uses a combination of physical and visual examinations of the breasts to check for signs and symptoms of breast cancer.

mammography: Mammography is an x-ray imaging method used to examine the breast for the early detection of cancer and other breast diseases.

breast cancer: Breast cancer is when breast cells mutate and become cancerous cells that multiply and form tumors.

## Data Availability

All data produced in the present study are available upon reasonable request to the authors

## Acknowledgements

Authors would like to thank the administration of Shri Atal Bihari Vajpayee Medical College and Research Institute for all the facilities provided for this study.

## Author contributions: CRediT

**Swastik Pandita:** Conceptualization, Methodology, Software, Writing-Reviewing and Editing **Andrea Dias**: Data curation, Writing-Original draft preparation, investigation. **Shirin Laturkar**: Visualization, Investigation. ***Shashank K A***: Supervision.

